# Coincidence of COVID-19 epidemic and olfactory dysfunction outbreak

**DOI:** 10.1101/2020.03.23.20041889

**Authors:** Seyed Hamidreza Bagheri, Alimohamad Asghari, Mohammad Farhadi, Ahmad Reza Shamshiri, Ali Kabir, Seyed Kamran Kamrava, Maryam Jalessi, Alireza Mohebbi, Rafieh Alizadeh, Ali Asghar Honarmand, Babak Ghalehbaghi, Alireza Salimi

## Abstract

**Background:** Recent surge of olfactory dysfunction in patients who were referred to ENT clinics and concurrent COVID-19epidemic in Iran motivated us to evaluate anosmic/hyposmic patients to find any relation between these two events.

**Methods:** This is a cross-sectional study with an online checklist on voluntary cases in all provinces of Iran between the 12th and 17th March, 2020. Cases was defined as self-reported anosmia/hyposmia in responders fewer than 4 weeks later (from start the of COVID-19 epidemic in Iran). Variables consist of clinical presentations, related past medical history, family history of recent respiratory tract infection and hospitalization.

**Results:** In this study 10069 participants aged 32.5±8.6 (7-78) years, 71.13% female and 81.68% non-smoker completed online checklist. They reported 10.55% a history of a trip out of home town and 1.1% hospitalization due to respiratory problems recently. From family members 12.17% had a history of severe respiratory disease in recent days and 48.23% had anosmia/hyposmia.

Correlation between the number of olfactory disorder and reported COVID-19 patients in all 31 provinces till 16th March 2020 was highly significant (Spearman correlation coefficient=0.87, p-Value<0.001). The onset of anosmia was sudden in 76.24% and till the time of filling the questionnaire in 60.90% of patients decreased sense of smell was constant. Also 83.38 of this patients had decreased taste sensation in association with anosmia.

**Conclusions:** It seems that we have a surge in outbreak of olfactory dysfunction happened in Iran during the COVID-19 epidemic. The exact mechanism of anosmia/hyposmia in COVID-19 patients’ needs further investigations.

## Introduction

Olfactory dysfunction following the upper respiratory tract infections also named post-viral anosmia has been reported in previous studies ^1,2^. Epithelial damage and central nervous system involvement, have been presented as the probable causes, however, the exact pathogenesis remains unclear ^2,3^. Generally, post-viral anosmia is more common in women and middle-aged or older individuals are more affected ^4^. Whilst the olfactory impairment can be permanent, this is often not the case and this kind of anosmia has more favorable prognosis than the other subgroups ^1^.

Coronaviruses are a large family of viruses that have been presented as the causative agent of different clinical manifestations ranging from a common cold to severe and great global public health concerns such as Middle East Respiratory Syndrome (MERS-CoV) and Severe Acute Respiratory Syndrome (SARS-CoV), so far; and corona virus disease 2019 (COVID-19) is a new strain from this family which has been introduced in the world since 2019 ^5,6^. The World Health Organization has reported 167515 confirmed cases of COVID-19 with 6606 deaths by March 16, 2020, globally ^7^. Because of the newly identified virus, it is expected that new reports about different aspects of the disease will be released daily ^6,8^; however, an update on its clinical and laboratory presentations has reported fever, respiratory symptoms, cough, fatigue, myalgia, arthralgia and breathing difficulties as the common presentations of the confirmed cases ^9^. To our knowledge, in none of the updates after the COVID-19 outbreak, anosmia is mentioned as one of the most prominent symptoms. Islamic Republic of Iran is one of the countries which has reported 14991 laboratory-confirmed cases of COVID-19 with 853 total deaths up to March 16, 2020 ^7^. Since no published scientific evidence reported olfactory and taste disorders following the COVID-19 pandemic till now; the aim of present study is an assessment of the frequency of olfactory disorder and patients’ characteristics in our country.

## Methods

This is a cross-sectional study on 10069 cases in all provinces of Iran between the 12th and 17th March, 2020. Participants were cases with problems in decreased sense of smell recently (the last month) invited to voluntarily respond to an online checklist, which was distributed in social networks. The online questionnaire developed by Ear, Nose, Throat and Head and Neck Surgery Research Center (Iran University of Medical Sciences) in cooperation with Iran Medical Council. The software incorporated into the website of Iran Medical Council for corona epidemics (www.corona.ir). The online questionnaire was in google document format (go.irimc.org/smelltest). The inclusion criteria were anosmia in responders fewer than 4 weeks later (from start the of COVID-19 epidemic).

The primary outcome of the study was anosmia-/-hyposmia of responders which measured by closed questions and scored by numerical scales at the commencement of their problem and its condition at the time of response to questionnaire. Secondary outcomes were clinical manifestations of participants such as flu or cold symptoms before anosmia, headache (which needed pain reliever drug), nasal stiffness, fever and chills, prominent cough, orbital (peri-orbital) pain, facial fullness and sinus pain, rhinorrhea, dyspnea, nasal irritation, parosmia, nasal pruritus, history of hypothyroidism, otalgia, sneezing (frequent), purulent nasal discharge, history of asthma, cheeks pain,, previous sinus surgery or septorhinoplasty, unilateral facial palsy and history of hypertension, diabetes mellitus and hyperthyroidism.

To check about validity of data, questions cross-validated with each other. For example, if a person says that they have hyposmia but in semi-quantitative assessment by our numerical scale pointed out the zero score, he/she was deleted from the analysis. To prevent data duplication, personal information of responders was compared in the analysis phase and to exclude incomplete responses to questionnaire the records with more than 30% missing data were deleted.

Categorical variables are reported as counts and percentages and quantitative variables as means and standard deviations (otherwise mentioned). Relationship between the number of participants and the number of confirmed COVID-19 patients (from national reports) analyzed by the Spearman correlation coefficient. This part of assessment has been considered as an ecological study because data are not at individual level. The study was approved by the ethics committee of Iran University of Medical Science. Filling online questionnaire considered as participants’ agreement with publishing the results in an anonymous way and in group (pooled) fashion.

## Results

During this period the online questionnaire viewed by 15228 people from who 10249 have had anosmia/hyposmia and answered the questions. Some participants’ information was excluded from analysis; 25 people who did not respond to more than 10 questions (out of 36 questions in the form), 155 records which were duplicated in personal information, 8 persons whose duration of disease was more than 30 days. So the final analysis is representative of 10069 responders. In demographic data; age distribution ranged from 7 to 78 years old (32.5±8.6), 71.13% were female and 81.68% non-smoker.

Most responders were from Gilan (51.9%), Tehran (18.4%), Mazandaran (6.6%), Golestan (4.3%) and Qom (2.7%) provinces. Despite a significant difference in the number of responders from multiple geographical areas, the distribution of demographic and clinical variables was homogenous among provinces of Iran.

Due to the COVID-19 epidemic in Iran, the number of cases of anosmia is compared to reported COVID-19 patients till 16th of March 2020 in figure-2. There is a significant correlation between anosmia and COVID-19 positivity in different provinces (Spearman correlation coefficient: 0.87, P-Value <0.001; figure-1)

**Figure-1.**
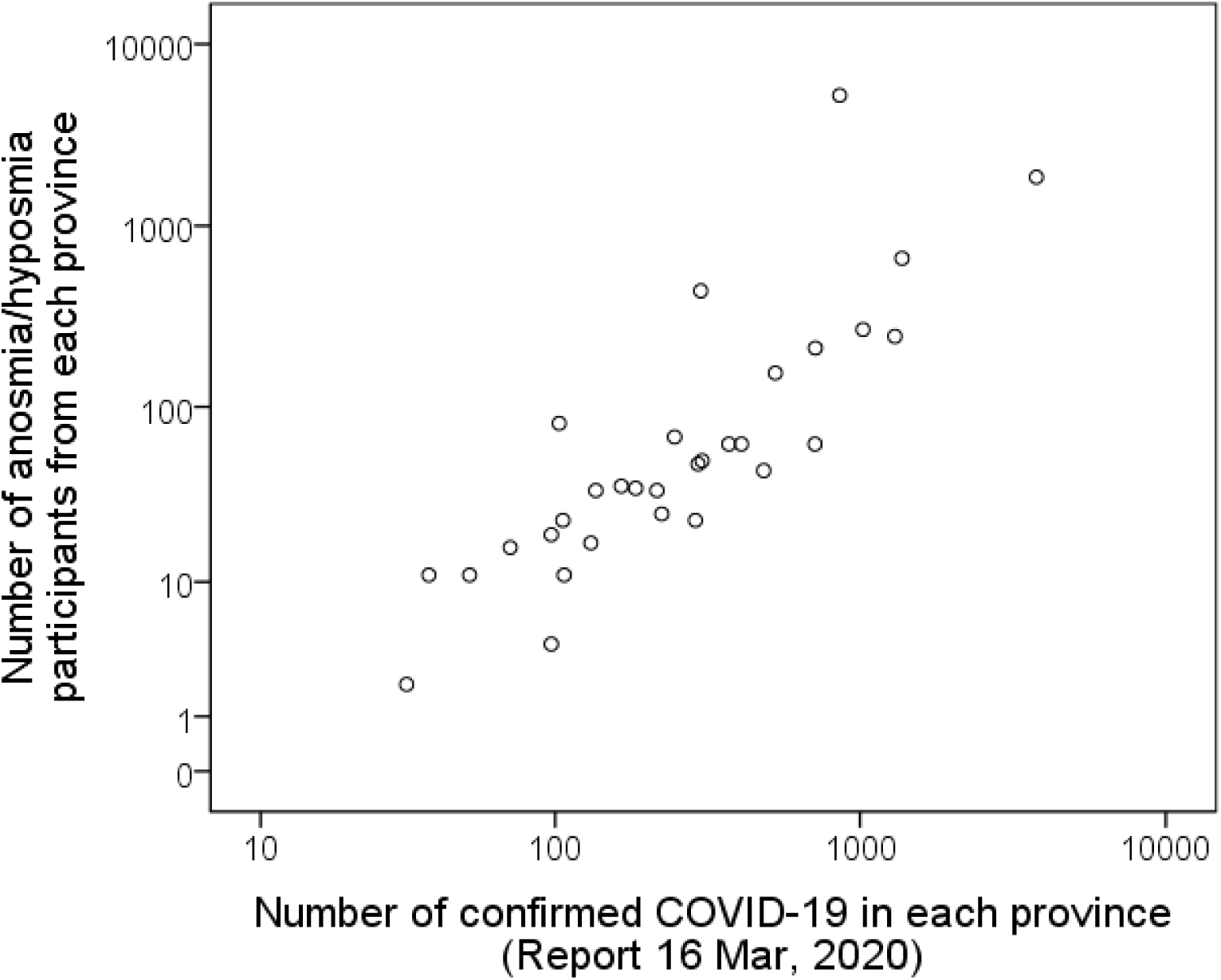
Correlation of anosmia and COVID-19 positivity in different provinces of Iran.

**Figure-2.**
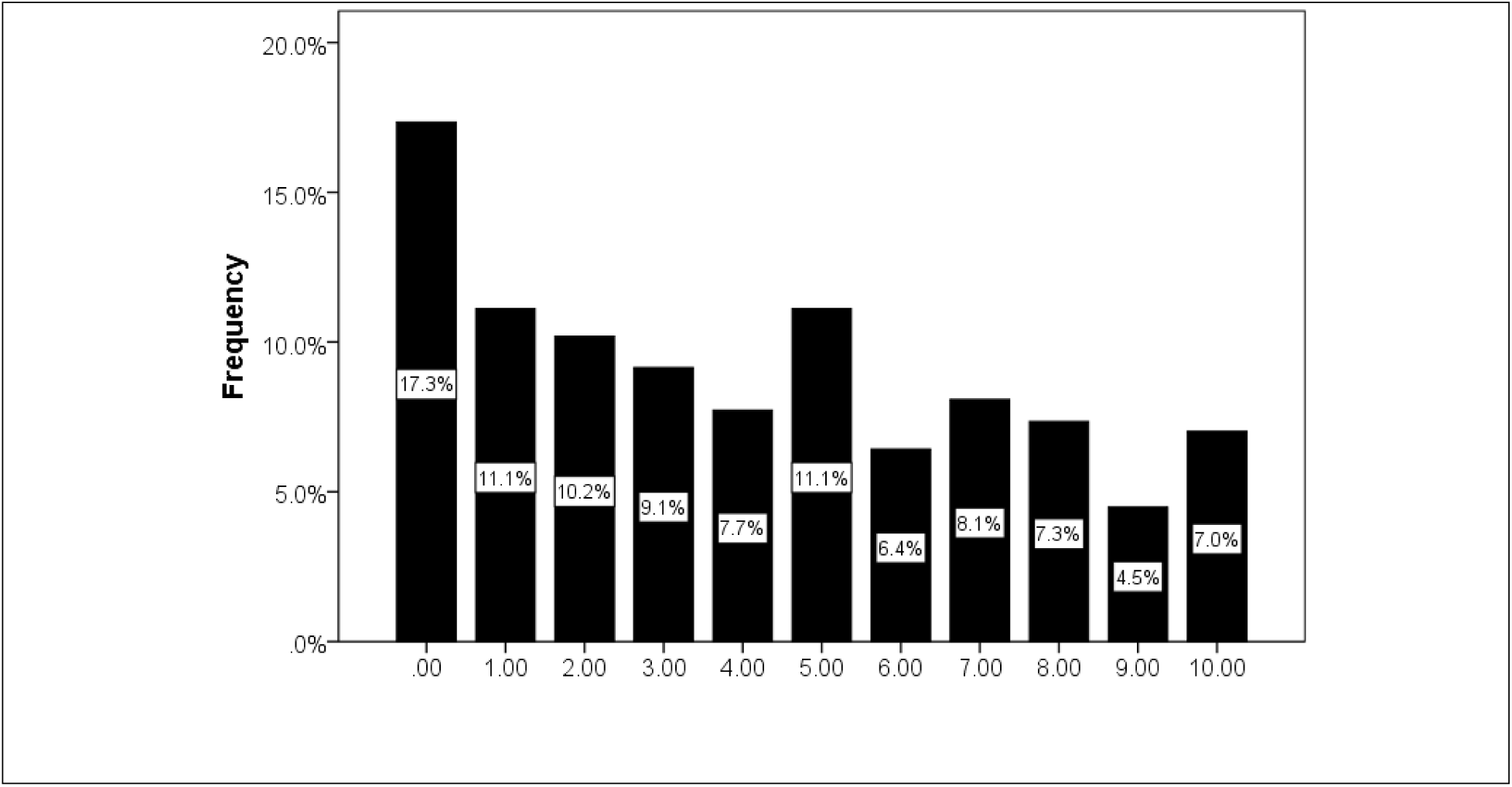
Numerical Scale of smell sensation at the time of filling questionnaire 0 (no sense of smell), 10 (full sense of smell)

From the clinical point of view, the onset of anosmia was sudden in 76.24% and till the time of filling the questionnaire in 60.90% of patients decreased sense of smell was constant. Also 83.38 of this patients had decreased taste sensation in association with anosmia. The other clinical findings are summarized in table-1.

**Table-1.** summary of clinical manifestations and past medical history of participants:

| Clinical manifestation | No. | % |
| --- | --- | --- |
| Flu or cold symptoms before anosmia | 7608 | 75.56% |
| Headache (which needed pain reliever drug) | 4903 | 48.69% |
| Nasal stiffness | 4400 | 43.70% |
| Fever and chills | 3764 | 37.38% |
| Sore throat | 1948 | 19.35% |
| Prominent cough | 1911 | 18.98% |
| Orbital (peri-orbital) pain | 1898 | 18.85% |
| Facial fullness and sinus pain | 1864 | 18.51% |
| Rhinorrhea | 1574 | 15.63% |
| Dyspnea | 1448 | 14.38% |
| Nasal irritation | 1380 | 13.71% |
| Parosmia | 1287 | 12.78% |
| Nasal pruritus | 1083 | 10.76% |
| Otalgia | 999 | 9.92% |
| Sneezing frequent | 914 | 9.08% |
| Purulent nasal discharge | 879 | 8.73% |
| Cheeks pain | 452 | 4.49% |
| Unilateral facial palsy | 52 | 0.52% |
| Past medical history |  |  |
| History of hypothyroidism | 1077 | 10.70% |
| History of asthma | 612 | 6.08% |
| History of hypertension | 428 | 4.25% |
| Previous sinus surgery or septorhinoplasty | 266 | 2.64% |
| History of diabetes mellitus | 199 | 1.98% |
| History of hyperthyroidism | 158 | 1.57% |
| History of epilepsy | 47 | 0.47% |

No.: number

Approximately, 10.55% of responders had a history of a trip out of home town before anosmia and about 1.1% of participants were hospitalized due to respiratory problems recently. From the family point of view; in 12.17% of responders one of the close relatives has had a history of severe respiratory disease in recent days, at least one of their family members was hospitalized due to respiratory problem in 7.35% of all and in 48.23% of them at least one of their family members has had recent history of anosmia.

At the time of response to questions the range of duration of anosmia onset was from 0 (the same day of onset) till 30 days (11.33±6.81, median=10.00) and more than 85% of responders did not use any drug for anosmia.

Use of a numerical scale which coded as 0 (no smell/taste, significantly decreased quality of life) to 10 (full smell/taste, normal quality of life) showed that significant number of patients had smell and taste problem at the time of filling the questionnaire (figures 2 and 3) and quality of life was obviously affected due to anosmia (figure 4).

**Figure-3.**
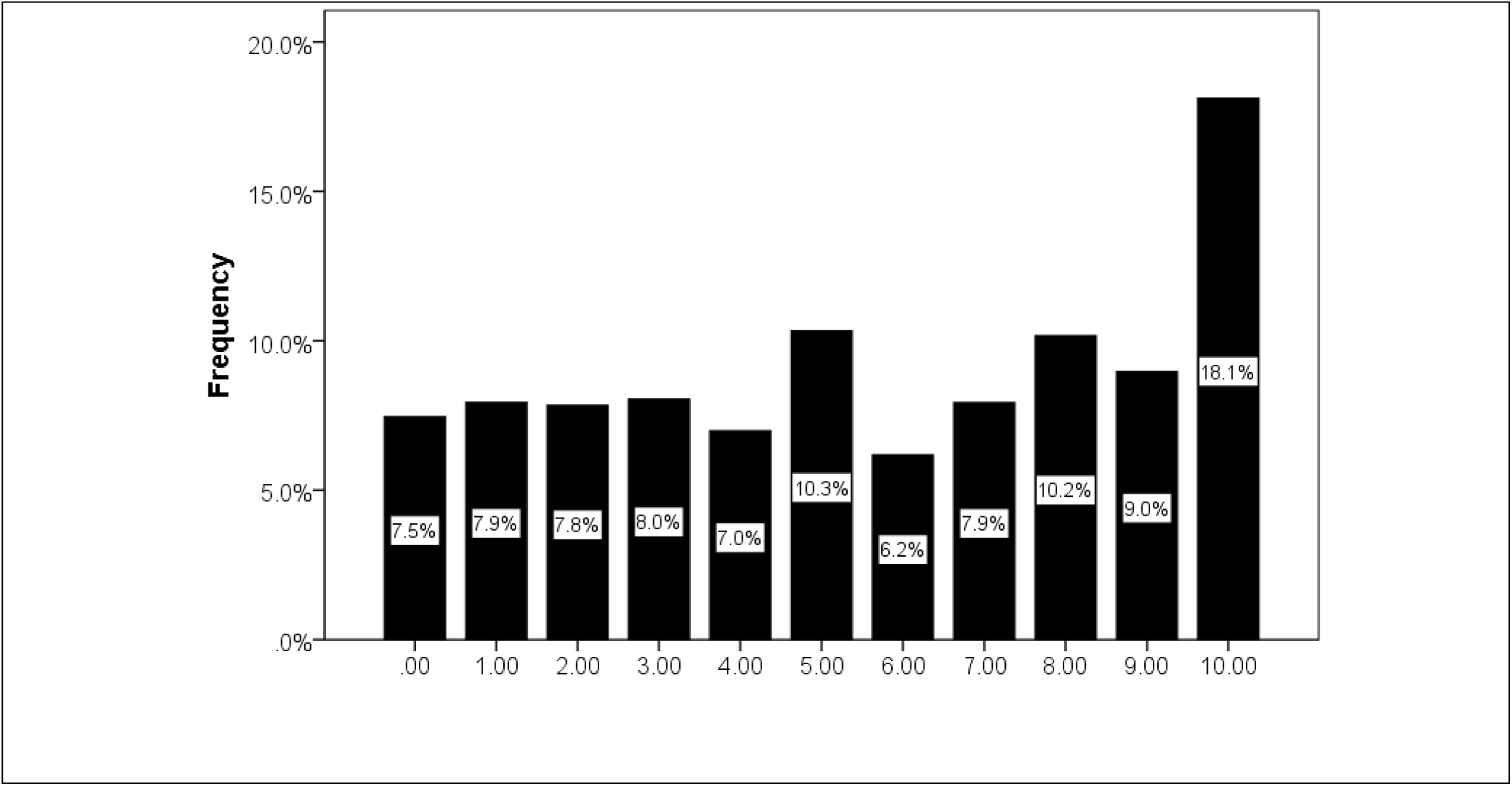
Numerical Scale of taste sensation at the time of filling questionnaire 0 (no sense of taste), 10 (full sense of taste)

**Figure-4.**
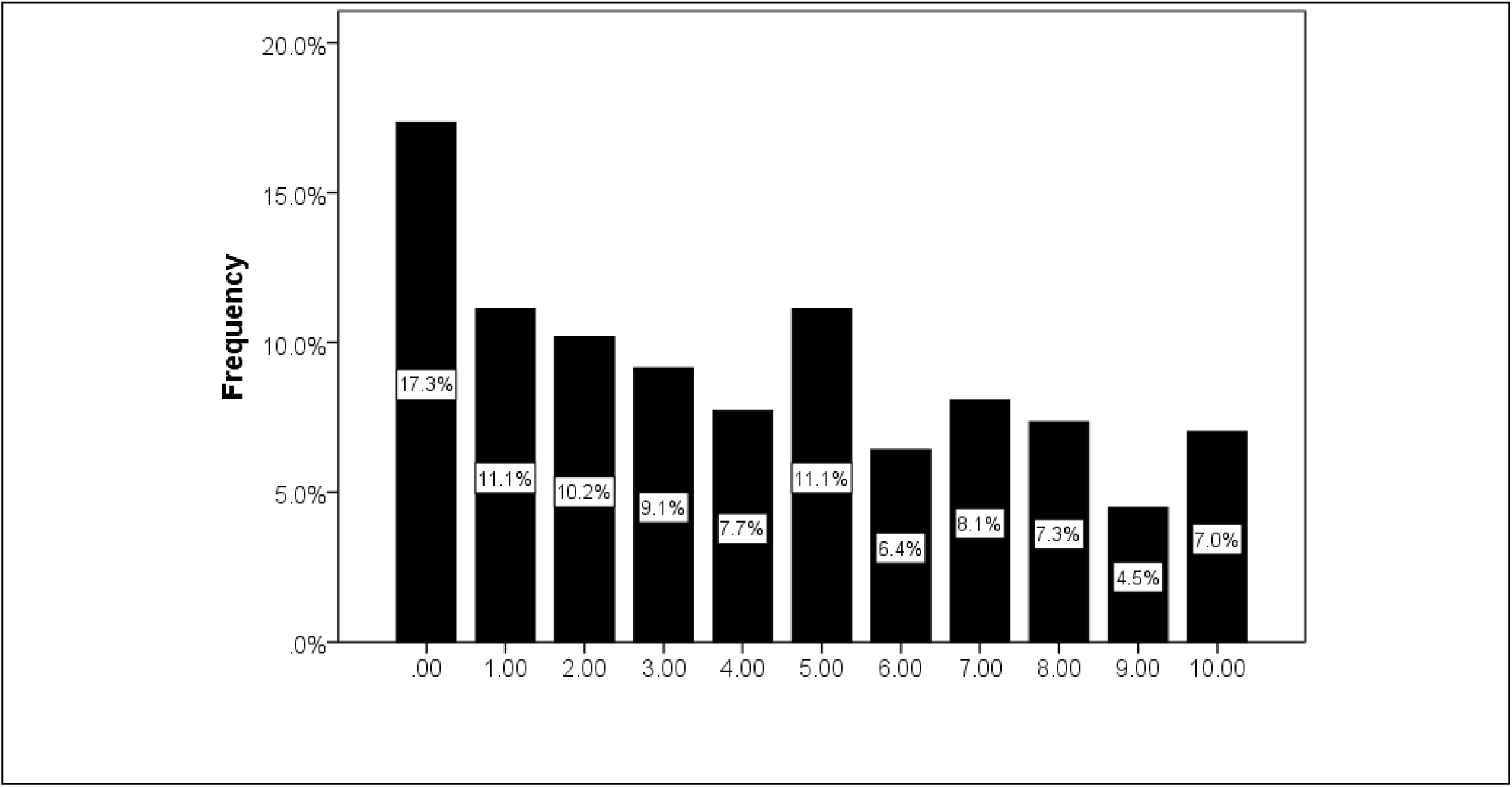
Numerical Scale of quality of life at the time of filling questionnaire 0 (significantly decreased quality of life), 10 (normal quality of life)

## Discussion

During the last month (March 2020), concurrent with COVID-19 epidemic, the number of patients with olfactory dysfunction was increased in different provinces of Iran. So we decided to assess the prevalence of this olfactory disorder by an online questionnaire. Till now results indicate the widespread prevalence of anosmia/hyposmia around the country and a significant linear correlation between prevalence of COVID-19 and olfactory impairment.

The recent great outbreak of olfactory dysfunction following flu like symptoms (75%), higher incidence in women (71%), and high incidence of anosmia in family members (48.23%) are suggestive of a post-viral epidemic olfactory dysfunction in Iran. Although our study could not confirm that COVID-19 is the definite cause of anosmia, but there is no other recent virus or flu epidemics in Iran except COVID-19. So, it seems that the recent anosmia outbreak in Iran is highly correlated to COVID-19.

Common COVID-19 symptoms such as fever, cough and dyspnea were less common in the participants with anosmia / hyposmia compliant (87.9% vs 37.38%, 67.7% vs 18.98% and 18.6% vs 14.38% respectively) and only about 1.1% of study population were hospitalized due to respiratory problems recently.

Studies of the prevalence of olfactory dysfunction in consecutive cases at olfactory clinics in the USA, Europe and Japan showed that the most common etiologies are: post-viral upper respiratory tract infections (18–45% of the clinical population) followed by nasal/sinus disease (7–56%), head trauma (8–20%), toxins/drugs (2–6%) and congenital loss (0–4%) ^10^. By now, we found no studies to report a high frequency of olfactory disorder coincidence with COVID-19 virus epidemic and this study could be the first report of an epidemic of COVID-19 with simultaneous olfactory dysfunction. In the study of Mao et al. 2020, detailed neurologic manifestations of the hospitalized patients with COVID-19 is reported which 5% of patients had hyposmia ^11^.

Suzuki et al. in 2007 detected rhinovirus, coronavirus, parainfluenza virus, and Epstein-Barr virus in the nasal secretion of post-viral olfactory disorders patients for the first time. Also, they suggested that rhinoviruses can target olfactory impairment through mechanisms other than nasal obstruction and that rhinoviruses could induce different time courses of olfactory dysfunction ^12^. Although the exact pathophysiology of post viral anosmia is unclear, but injury could probably happen at the level of the neuro-epithelium of olfactory receptor cells in the nasal roof or in central olfactory routs, however, because of the accessibility of the olfactory cleft, many studies have focused on the neuro-epithelial changes in patients with post-viral olfactory dysfunction. Various animal studies have explained that different viruses could damage central olfactory routs and other regions of the brain ^13-16^. The neurological manifestations of COVID-19 have not reported in many previous studies yet, Mao et al. (2020) Showed that 36.4% of patients with COVID-19 had a variety of neurological symptoms involved CNS, PNS and skeletal muscles. CNS findings were the significant form of neurological damage in patients with COVID-19 in their study ^11^. The movement of the COVID-19 virus into the brain through the cribriform plate near to the olfactory bulb could be a route that would enable the virus to enter and influence the brain. Furthermore, the findings such as anosmia or hyposmia in an uncomplicated early stage COVID-19 patient should necessitate a thorough assessment for CNS involvement ^17^.

Our data is collected via an online self-reported questionnaire. The responders were part of the population who are literate and have access to the internet, personal computers or smart-phones. We announced the patients to answer this questionnaire via social networks so the participants are limited to those who use social networks. However, because of high prevalence of using smart phone even by housekeepers and people with education level lower than diploma, representativeness of the results does not seem to be affected by this bias. Another limitation of this study is that no definite test for COVID-19 infection was included. As this is an online survey, the main goal was to confirm the outbreak of anosmia during the COVID-19 epidemic. As the study was self-reported, we used some of questions to check others to increase the questionnaire response validity.

Based on the findings of this study it seems that we have a surge in outbreak of olfactory dysfunction happened in Iran during the COVID-19 epidemic, that correlates with the number of patients infected with COVID-19 across the country. As olfactory dysfunction can affect the quality of life in affected patients, it needs to be assessed worldwide by further clinical studies to find out the exact correlation, pathogenesis, prognosis, and any correlation between disease severity and olfactory dysfunction.

## Data Availability

The online questionnaire developed by Ear, Nose, Throat and Head and Neck Surgery Research Center (Iran University of Medical Sciences) in cooperation with Iran Medical Council. The software incorporated into the website of Iran Medical Council for corona epidemics (www.corona.ir). The online questionnaire was in google document format (go.irimc.org/smelltest).

## References

1. Hummel T, Whitcroft K, Andrews P, et al. Position paper on olfactory dysfunction. Rhinology Supplement 2017;54.

2. Welge-Lüssen A. Re-establishment of olfactory and taste functions. GMS current topics in otorhinolaryngology, head and neck surgery 2005;4.

3. Hummel T, Landis BN, Hüttenbrink K-B. Smell and taste disorders. GMS current topics in otorhinolaryngology, head and neck surgery 2011;10.

4. Jafek BW, Murrow B, Michaels R, Restrepo D, Linschoten M. Biopsies of human olfactory epithelium. Chemical senses 2002;27:623–8.

5. World Health Organization, 2020 March 2020; Available from: https://www.who.int/health-topics/coronavirus. 2020.

6. Li G, De Clercq E. Therapeutic options for the 2019 novel coronavirus (2019-nCoV). Nature Publishing Group; 2020.

7. Organization WH. Coronavirus disease 2019 (COVID-19): situation report, 45. 2020.

8. Rothan HA, Byrareddy SN. The epidemiology and pathogenesis of coronavirus disease (COVID-19) outbreak. Journal of Autoimmunity 2020:102433.

9. Chan KW, Wong VT, Tang SCW. COVID-19: An Update on the Epidemiological, Clinical, Preventive and Therapeutic Evidence and Guidelines of Integrative Chinese–Western Medicine for the Management of 2019 Novel Coronavirus Disease. The American Journal of Chinese Medicine 2020:1–26.

10. Nordin S, Brämerson A. Complaints of olfactory disorders: epidemiology, assessment and clinical implications. Current opinion in allergy and clinical immunology 2008;8:10–5.

11. Mao L, Wang M, Chen S, et al. Neurological Manifestations of Hospitalized Patients with COVID-19 in Wuhan, China: a retrospective case series study. 2020.

12. Suzuki M, Saito K, Min WP, et al. Identification of viruses in patients with postviral olfactory dysfunction. The Laryngoscope 2007;117:272–7.

13. Seiden AM. Postviral olfactory loss. Otolaryngologic Clinics of North America 2004;37:1159–66.

14. Duncan H. Postviral olfactory loss. Taste and smell disorders 1997:172–8.

15. Mohammed A, Magnusson O, Maehlen J, et al. Behavioural deficits and serotonin depletion in adult rats after transient infant nasal viral infection. Neuroscience 1990;35:355–63.

16. Perlman S, Evans G, Afifi A. Effect of olfactory bulb ablation on spread of a neurotropic coronavirus into the mouse brain. The Journal of experimental medicine 1990;172:1127–32.

17. Baig AM, Khaleeq A, Ali U, Syeda H. Evidence of the COVID-19 Virus Targeting the CNS: Tissue Distribution, Host–Virus Interaction, and Proposed Neurotropic Mechanisms. ACS Chemical Neuroscience 2020.

